# Effectiveness of pharmacy-based HIV pre- and post-exposure prophylaxis delivery: a cluster-randomized trial in Kenya

**DOI:** 10.64898/2026.06.26.26356703

**Authors:** Katrina F. Ortblad, Allison Meisner, Victor Omollo, Tabitha Kareithi, Stephanie D. Roche, Patricia Ong’wen, Magdaline Asewe, Micah Onenga Anyona, Preetika Banerjee, Kelly Curran, Eunice Gichuru, Kendall Harkey, Lawerence Juma, Catherine Kiptinness, Rachel C. Malen, Melissa L. Mugambi, Peris Otieno, Jillian Pintye, Bernard Rono, Torin T. Schaafsma, Parth D. Shah, Monisha Sharma, Katherine K. Thomas, Kaiyue Yu, Daniel Were, Elizabeth A. Bukusi, Kenneth Ngure, the Pharm PrEP study team

## Abstract

Private pharmacies are ubiquitous yet underutilized for HIV pre- and post-exposure prophylaxis (PrEP and PEP) delivery. In a cluster-randomized trial in Kenya (NCT05842122), we randomized 60 pharmacies 1:1:1:1 to: client-sustained delivery (∼$2/visit user fee); implementor-sustained delivery (∼$2/visit reimbursement); counselor-supported delivery (task shifting; ∼$1/visit reimbursement); or clinic referral (control; ∼$1/referral reimbursement). Commodities were supplied free to pharmacies from government stock. Primary outcomes were PrEP initiation and one-month continuation (any dispensing or refilling, respectively), self-reported by clients 60 days post-enrollment (multiple-comparisons threshold: p=0.017). From June 2023-April 2025, 5,808 clients enrolled; 64% were PEP candidates. Compared to referral, the counselor-supported arm had significantly higher PrEP initiation (RR=6.5, 95% CI [2.6, 16], p<0.001) and continuation rates (RR=5.1, 95% CI [1.4, 19], p=0.016); all intervention arms had significantly higher PEP initiation rates. One seroconversion, one social harm, and two provider needlestick injuries occurred. Pharmacy PrEP/PEP delivery outperformed clinic referral, particularly when fully subsidized and counselor-supported.

## INTRODUCTION

Private pharmacies are a promising yet underleveraged platform for HIV prevention service delivery. Compared to public health facilities, pharmacies offer greater accessibility through convenient locations, extended operating hours, and rapid, discreet services.^1,2^ For these reasons, they are frequently accessed for sexual and reproductive health (SRH) needs—including contraception, sexually transmitted infection (STI) treatment, and sexual performance enhancement—making them a convenient entry point for HIV prevention.^2-4^ Global and national policymakers, including the Kenya Ministry of Health (MOH), have increasingly recognized this potential and are prioritizing pharmacy-delivered services in HIV programs.^5,6^ However, gaps in the policy and regulatory environment continue to constrain implementation, and evidence on effective implementation strategies to support delivery at scale is lacking.

We developed one of the first pharmacy provider-led models of HIV pre- and post-exposure prophylaxis (PrEP and PEP) initiation and management in Africa. Adapted from a Seattle-based collaborative practice agreement model^7^ and refined for Kenya though formative research,^1^ multi-stakeholder consultation,^8^ and several pilot studies,^9,10^ the model enables licensed pharmacist and pharmaceutical technologists to initiate and manage clients on PrEP and PEP using a prescribing checklist, under remote oversight by prescribing physicians and clinical officers.^11^ Pilot studies demonstrated the model’s feasibility, acceptability, and reach to individuals underrepresented at public clinics, including unmarried individuals and those <25 years.^9,11-13^ Findings also indicated client’s willingness to pay out of pocket for pharmacy PEP/PrEP and delivery costs comparable to facility-based care.^14,15^ However, scale-up remains constrained by policy gaps (e.g., limitations on pharmacy provider scope of practice; exclusion of pharmacies from the national PrEP/PEP supply chain), limited evidence on sustainable financing approaches, and misalignment between PrEP/PEP delivery’s time-intensive nature (∼40 minutes/visit in pilot studies)^14^ and private pharmacy revenue models.

To inform policy and scale-up strategies, we conducted a four-arm cluster-randomized controlled trial (cRCT) comparing three pharmacy-based PrEP/PEP “direct delivery” models to an active comparator: pharmacy-based PEP/PrEP eligibility screening with referral to clinic-based services, a feasible standard under existing Kenya guidelines.^5^ The first direct delivery model aligned with the approach outlined in Kenya’s Private Sector Engagement Framework:^6^ a partial subsidy model in which commodities are supplied at no cost from linked public clinics, and clients pay a user fee for services. The remaining two models tested additional layered implementation strategies: a full subsidy model (no out-of-pocket client costs; user fees covered by the implementer) and task-shifting of time-intensive HIV testing and counseling steps^14^ to a dedicated HIV testing services (HTS) counselor. This trial provides the first randomized evidence on the comparative effectiveness of pharmacy-based PrEP/PEP delivery models compared to clinic referral.

## METHODS

### Study design and setting

The Pharm PrEP cRCT (NCT04982250) was a four-arm, cluster-level, parallel trial conducted in six counties across two regions of Kenya—Western (Kisumu, Siaya, Migori, Homa Bay) and Central (Nairobi, Kiambu)—selected to capture regional variation in HIV epidemiology and PrEP context. Western Kenya has substantially higher population-level HIV prevalence than Central Kenya (∼11% vs. ∼2%)^16^ and greater PrEP awareness due to longstanding education campaigns and implementation efforts, as well as lower average household income. Details on study procedures have been published elsewhere.^17^

### Study pharmacies

We collaborated with county and sub-county health management teams to identify private retail pharmacies meeting our eligibility criteria: registration with Kenya’s Pharmacy and Poisons Board, a full-time licensed pharmacist or pharmaceutical technologist on staff, a private room for HIV testing and counseling, and willingness to participate in research activities. To minimize contamination risk between arms, study pharmacies had to be at least three kilometers apart. In total, 80 pharmacies were selected for training and randomization: 60 study and 20 back-up pharmacies.

Up to two providers per pharmacy completed a three-component training comprising a self-paced online course, two-day in-person course, and observed HIV testing practice session overseen by an MOH trainer. We drew course content from existing MOH curricula on HIV rapid diagnostic testing (RDT) and daily oral PrEP and PEP delivery,^18,19^ tailoring materials to align with study procedures. Throughout implementation, study staff with clinical training delivered routine technical assistance—more intensively during start-up and tapering thereafter—and conducted on-site refresher trainings, as needed (e.g., following pharmacy provider turnover).

### Study participants

Eligible pharmacy clients were ≥16 years, interested in PrEP or PEP screening, and medically eligible per the prescribing checklist.^17^ Recruitment occurred through provider-initiated conversations with clients purchasing SRH products, in-pharmacy posters, and unrestricted pharmacy-chosen demand generation strategies.

The study protocol was approved by the Kenya Medical Research Institute’s Scientific Ethics Review Unit and Fred Hutchinson Cancer Center’s Institutional Review Board (**Supplementary File 1. Protocol**). After pharmacy providers conducted PEP/PrEP eligibility assessments, an off-site research assistant (RA) obtained verbal consent by phone to minimize influence on service delivery and replicate real-world implementation conditions. Participants received no compensation for enrolling but were given 250 KES (∼$2 USD) for completing a 60-day survey. Pharmacies received 200 KES (∼$1.50 USD) monthly as transport reimbursement for collecting commodities from linked public clinics. A Data Safety and Monitoring Board (DSMB) convened every six months to review progress, safety outcomes, and offer guidance.

### Randomization and masking

We categorized study pharmacies into four geographic groups—Kisumu/Siaya and Migori/Homa Bay Counties in Western Kenya, and Kiambu and Nairobi Counties in Central Kenya—and used stratified randomization to assign study arms. The 20 pharmacies in each group were randomized to one of four study arms (15 pharmacies, 1:1:1:1 allocation) or a backup pool (five pharmacies; see **Supplementary File 2. Statistical Analysis Plan** for details). Following in-person training, one provider per pharmacy drew an opaque sealed envelope containing their arm assignment or backup designation. Allocation sequences were generated in advance by the study biostatistician (author AM) independent of pharmacy selection and enrollment. Arm assignment was unblinded due to the nature of the interventions. Pharmacies that withdrew or became ineligible were replaced by random selection from the backup pool.

### Procedures

Providers at all study pharmacies used an electronic version of the prescribing checklist^8,17^—programmed into a point-of-sale (POS) platform (Maisha Meds, Kisumu, Kenya)—to screen clients for PrEP/PEP eligibility in a private room. The checklist assessed ongoing HIV risk, using a modified 13-item version of Kenya’s HIV Risk Assessment Screening Tool, which asked clients to self-report select behaviors (e.g., transactional sex) in the past 6 months;^20^ acute HIV risk, via three items on potential exposures (condomless sex, shared needles, sexual assault) in the past 72 hours; and PrEP medical safety, by screening for signs of acute HIV infection (e.g., swollen lymph nodes) and history of liver or kidney disease, diabetes, or hypertension. Clients with no HIV risk indication were ineligible for enrollment, as were those reporting medical conditions that could contraindicate PrEP; these clients were referred to a clinic of their choice for further evaluation. Pharmacy providers determined preliminary PEP or PrEP candidacy following counseling and prior to HIV testing; a remote study clinician was available 24/7 for consultation.

Pharmacy providers in direct delivery pharmacies confirmed HIV-negative status using RDT and dispensed PrEP or PEP to eligible clients without creatinine or hepatitis B/C testing, consistent with national guidelines permitting initiation absent these tests.^21^ Clients who tested HIV-negative were dispensed a 30-day supply of daily oral PrEP (tenofovir disoproxil fumarate with emtricitabine or lamivudine) or 28-day supply of daily oral PEP (tenofovir disoproxil fumarate/lamivudine or emtricitabine/dolutegravir) and were scheduled for a follow-up visit one month later; clients testing HIV-positive were referred to nearby clinics for confirmatory testing and treatment. At follow-up, clients were screened for PrEP/PEP side effects (e.g., headache, nausea) and acute HIV infection symptoms, and underwent HIV testing. PEP clients who tested HIV-negative were offered PrEP eligibility screening; PrEP clients who elected to continue and met eligibility criteria were dispensed a 90-day supply. Follow-up visit reminders were left to each pharmacy’s existing practices (e.g., phone calls, SMS messages).

All direct delivery pharmacies operated under a mixed financing model reflective of a public-private partnership, in which government-supplied PrEP/PEP drugs and HIV testing kits—provided to pharmacies at no cost from linked public clinics—constituted an in-kind commodity subsidy. To cover remaining costs associated with provider time and services, a partial subsidy arm applied a user fee (cost-sharing with clients), while two full subsidy arms eliminated client cost-sharing through per-visit financial reimbursements to pharmacies—analogous to provider reimbursement models in which a public-sector payer or private insurer covers the full cost of care. One of the full subsidy arms additionally employed a dedicated HTS counselor to task-shift the time-intensive HIV testing and counseling steps^14^ from the pharmacy provider. User fees and reimbursement amounts were informed by pilot studies assessing client willingness to pay and provider willingness to charge,^14^ as well as consultations with pharmacy providers and government officials. The differences between the direct delivery arms and clinic referral arm are described in detail below.

1. <u>Client-sustained delivery</u>: Clients paid a user fee of 250 Kenyan Shillings (KES; ∼$2 US Dollars [USD]) per PrEP/PEP visit; pharmacy providers completed all delivery steps.
2. <u>Implementor-sustained delivery</u>: The study reimbursed pharmacies 250 KES per PrEP/PEP visit; clients paid nothing. Pharmacy providers completed all delivery steps.
3. <u>Counselor-supported delivery</u>: The study paid for and stationed an HTS counselor at pharmacies on weekdays from 9am to 5pm to complete HIV testing and counseling; the pharmacy provider conducted the medical safety assessment, offered additional counseling as needed, dispensed PrEP or PEP, and scheduled follow-up. The study reimbursed pharmacies 100 KES (∼$1 USD) per visit, reflecting providers’ reduced workload relative to the other direct delivery arms; clients paid nothing.
4. <u>Clinic referral (control</u>): Pharmacy providers screened clients for preliminary PEP or PrEP eligibility—stopping short of HIV testing—and referred interested candidates to a nearby clinic of the client’s choosing. The study reimbursed pharmacies 100 KES per referral.

### Data collection

We captured pharmacy and participant client characteristics and outcomes across four data sources: pharmacy surveys, pharmacy client volume data, the prescribing checklist, and client surveys.

Baseline pharmacy surveys, completed by pharmacy owners, captured pharmacy characteristics such as location, staffing, service offering, and clientele. We estimated pharmacies’ monthly client volume—a key statistical model input—by providing pharmacies with bags to use for all sales over one month and tracked bags used as a proxy for client volume. To account for season variation, we repeated this tracking activity three times: in July 2024, November 2024, and March 2025. This practical approach mitigated risk of inaccuracies arising from variation in pharmacy record-keeping practices (e.g., paper logs versus electronic POS systems) and record completeness.

The prescribing checklist captured participant characteristics at enrollment, including self-reported behaviors associated with HIV risk, and—for direct delivery arms—HIV test results and PrEP/PEP dispensing information. Client surveys assessed our primary (PrEP) and secondary (PEP and PrEP/PEP) outcomes consistently across all arms, including the clinic referral arm, for which dispensing data were unavailable. Surveys were conducted at 60 days post-enrollment to allow sufficient time to assess linkage to PrEP/PEP in the clinic referral arm and one-month PrEP continuation across all arms. Trained RAs administered surveys by phone and entered data into REDCap (v17.0.5).^22-25^ Participants not reached after three contact attempts were classified as lost to follow-up.

### Outcomes

Clients self-reported all outcomes at 60 days following enrollment. Primary outcomes were PrEP initiation (any PrEP dispensing at a pharmacy or clinic within this window) and one-month PrEP continuation (a PrEP refill following initiation, within this same window). Secondary outcomes, assessed within the same window, included: PEP initiation (any PEP dispensing), recurrent PEP use (PEP dispensing more than once), PEP-to-PrEP transition (PrEP dispensing following PEP initiation), PrEP/PEP initiation (dispensing of either product), and PrEP/PEP continuation (refill or dispensing of either product following initiation). We report implementation outcomes (e.g., acceptability, fidelity, costs) elsewhere.^15,26^

### Power

We powered the trial on pharmacy-level count outcomes for PrEP initiation and continuation. Assumed monthly initiation and continuation counts for the client-sustained and implementor-sustained arms were informed by two pharmacy-based pilot studies;^9,11^ those for the clinic referral arm were informed by a large clinic-based PrEP implementation project.^27^ Compared to clinic referral, we hypothesized PrEP initiation would be highest in the counselor-supported arm (∼2.6x), followed by the implementor-sustained (∼2.2x) and client-sustained arms (∼1.6x); we hypothesized PrEP continuation among initiators of 80%, 70%, 50%, and 60% in the counselor-supported, implementor-sustained, client-sustained, and clinical referral arms, respectively. The trial was originally powered using individual-level binary outcomes among enrolled participants purchasing SRH products; following substantially differential enrollment across arms early in implementation, the power calculation was amended to pharmacy-level count outcomes following DSMB approval (**Supplementary File 2. Statistical Analysis Plan**).

Power estimates assumed a coefficient of variation of 0.25 (typical for cRCTs), a Bonferroni-adjusted two-sided significance level of 0.017 to account for three primary comparisons (each direct delivery arm versus clinic referral), and equivalent average pharmacy client volume across arms. We estimated the number of PrEP initiations needed to detect the smallest hypothesized difference—between the client-sustained and the clinic referral arms—at 80% power, yielding 1,119 PrEP initiations across arms. This sample provides >80% power for all remaining direct delivery versus clinic referral comparisons, and for all PrEP continuation comparisons except the client-sustained versus clinic referral comparison (23% power).

### Statistical analysis

All analyses were intention-to-treat and included data from all pharmacies, including dropped and replaced pharmacies, during the period they were enrolled. For all outcomes, we conducted a pharmacy-level analysis of counts, accounting for pharmacy client volume and months enrolled (including implementation pauses), deviating from our originally planned proportions-based analysis. We imputed missing pharmacy client volume estimates using pharmacy-level characteristics and averaged values across three observation points. We present outcome counts by arm alongside annual pharmacy-level rates standardized to the median client volume of clinic-referral arm pharmacies (2,030 clients/month) to enable comparisons across pharmacies of varying size; standardized rates are for descriptive purposes only.

We estimated rate ratios (RRs) comparing each direct delivery arm to clinic referral using a Poisson generalized linear model with a log link and robust standard errors.^28^ The model included an offset for total pharmacy client volume—each pharmacy’s average monthly client volume multiplied by the number of months enrolled—and was adjusted for county group to reflect the stratified randomization. All statistical tests were two-sided, with estimated 95% confidence intervals (CIs). We applied a Bonferroni-adjusted significance threshold of p<0.017 to our primary PrEP and secondary PrEP/PEP outcomes, and p<0.05 to all remaining outcomes. Missing outcomes were imputed using observed pharmacy-specific outcome proportions, deviating from our pre-specified assumption that missing client survey responses represented failures, as pharmacy dispensing records indicated a sizable number of initiations among clients with missing survey data.

Pre-specified sensitivity analyses for primary outcomes included incorporating pharmacy-level implementation pauses and a non-parametric permutation test. We conducted a post-hoc sensitivity analysis excluding high-volume pharmacies (>5,000 clients/month) after observing that no counselor-supported pharmacies exceeded this threshold. A pre-specified secondary analysis estimated differences in the proportion of PrEP initiators continuing PrEP in each direct delivery arm compared to clinical referral, and subgroup analyses examined outcomes by region (Western versus Central Kenya) and pharmacy setting (urban versus rural/peri-urban). We additionally conducted all primary and secondary analyses under our original assumption that unobserved outcomes represented failures.

To understand pharmacy-specific performance, we plotted each pharmacy’s mean standardized annual rates of PrEP initiation, PrEP continuation, and PEP initiation by arm, scaling marker size to total pharmacy client volume over the implementation period.

We conducted all analyses in R version 4.6.0 (R Core Team, 2026).^29^

## RESULTS

### Pharmacies

From November 22, 2022 to March 17, 2023, we screened 411 pharmacies and initially randomized 60, 15 per arm (client-sustained, implementor-sustained, counselor-supported, and clinic referral; **Figure 1**). Among 336 ineligible pharmacies, primary exclusion reasons were lack of owner interest or reachability (43%, k=143), absence of a private room (10%, k=35), and lack of a full-time licensed pharmacist or pharmaceutical technologist (8.6%, k=29). Over the implementation period, 15 pharmacies were dropped and replaced, yielding 75 total study pharmacies: 20 client-sustained, 17 implementor-sustained, 18 counselor-supported, and 20 clinic referral pharmacies. Primary replacement reasons were provider turnover without timely replacement (60%, k=9) and prolonged provider absence (20%, k=3). Across all pharmacies, the median monthly client volume was 2,383 (interquartile range [IQR] 1,470, 3,942) and months enrolled was 21.1 (IQR 11.9, 21.5), with some variation by arm (**Supplementary Figure 2**). Study pharmacies were widely distributed across the four geographic groups (**Figure 2**).

**Figure 1.**
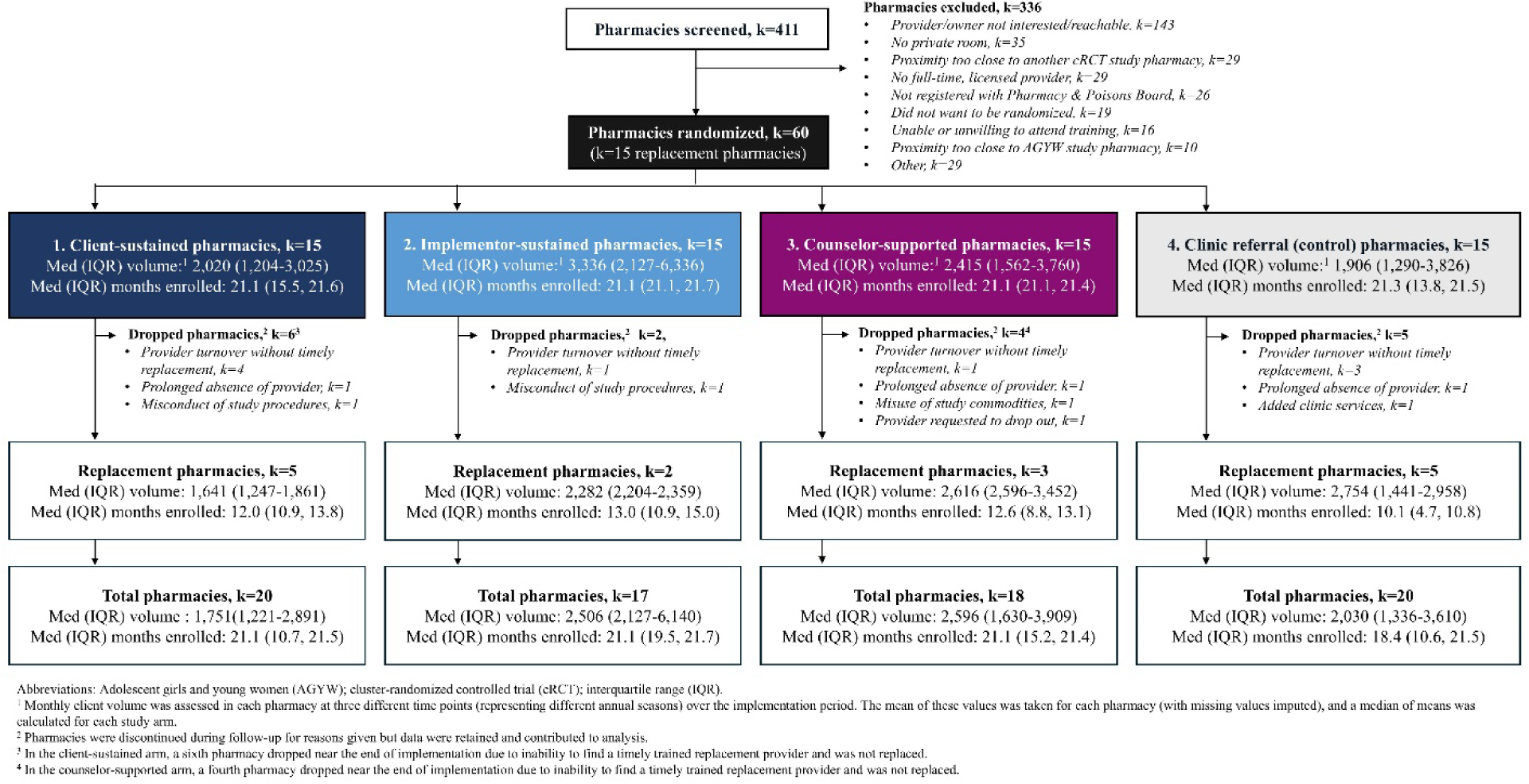
CONSORT diagram.

**Figure 2.**
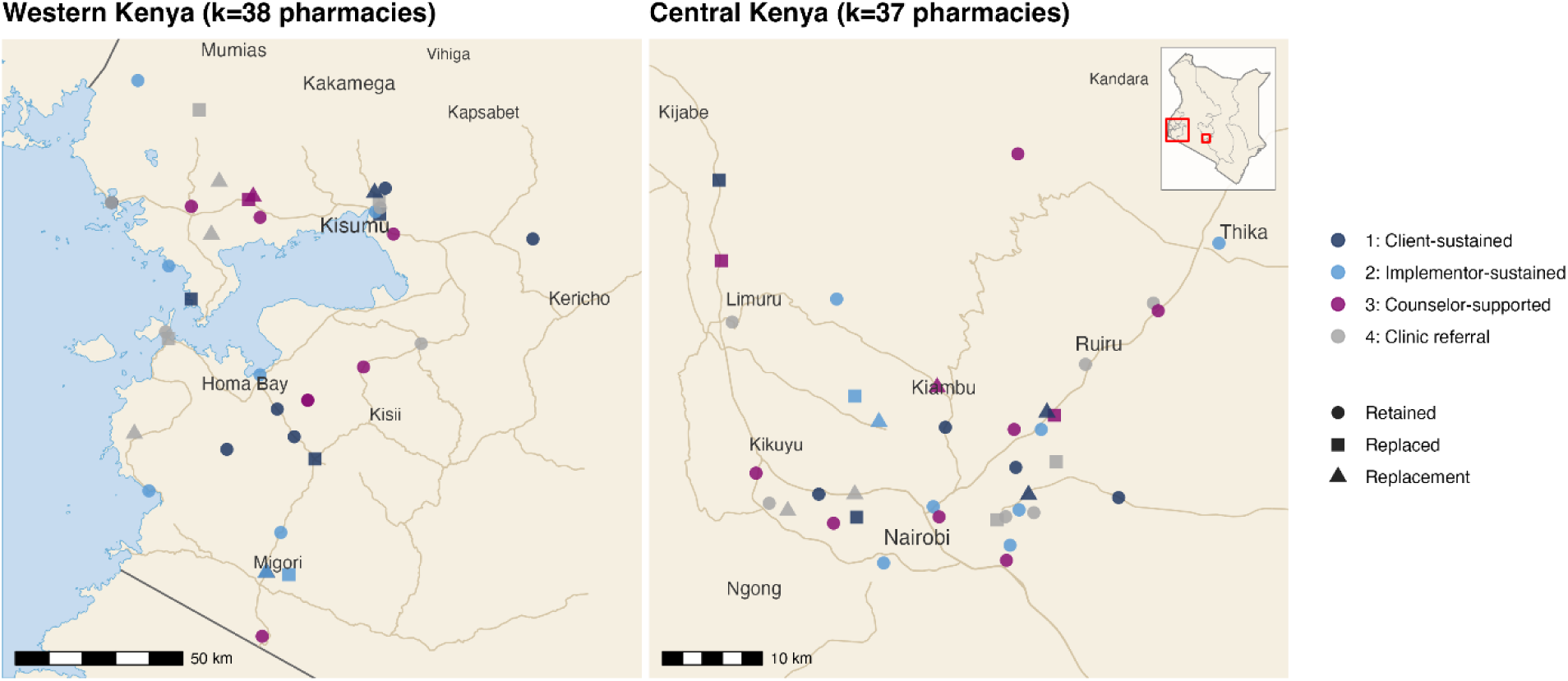
Map of study pharmacies included in our analysis.

Characteristics of the 75 study pharmacies are described in **Table 1**. Most were in urban settings (73%, k=55), with nearly half in middle- or high-income areas (46%, k=35). Many were located near markets (83%, k=62) or bars and night clubs (75%, k=56) and within 15 minutes walking of a public healthcare facility (68%, k=51). Most were independently owned (80%, k=60) with a median of 2 permanent staff (IQR 1, 2). Prior to study participation, most sold HIV self-testing kits (92%, k=69), some offered RDT services (23%, k=17), and none provided HIV prevention or treatment drugs. Roughly half offered follow-up reminders (43%, k=32), primarily via phone call (35%, k=26).

**Table 1.** Characteristics of study pharmacies (k=75), by arm.

| Characteristics<sup>1</sup> | 1: Client-sustained,<br>k=20 | 2: Implementor-sustained,<br>k=17 | 3: Counselor-supported,<br>k=18 | 4: Clinic referral (control),<br>k=20 |
| --- | --- | --- | --- | --- |
| **Pharmacy size** |  |  |  |  |
| Pharmacy area, m<sup>2</sup> (Median, IQR) | 32 (23–37) | 32 (15–48) | 31 (21–54) | 37 (19–48) |
| Consultation room area, m<sup>2</sup> (Median, IQR) | 12 (12–16) | 15 (10–24) | 15 (8–20) | 15 (11–17) |
| **Pharmacy setting** |  |  |  |  |
| Pharmacy location |  |  |  |  |
| Urban, middle/high income | 10 (50%) | 7 (41%) | 8 (44%) | 10 (50%) |
| Urban, lower income/informal settlement | 4 (20%) | 5 (29%) | 3 (17%) | 3 (15%) |
| Urban, Central Business District | 3 (15%) | 1 (6%) | 1 (6%) | 0 (0%) |
| Sub-urban/Peri-urban | 1 (5%) | 2 (12%) | 5 (28%) | 3 (15%) |
| Rural | 2 (10%) | 2 (12%) | 1 (6%) | 4 (20%) |
| Venues frequented by pharmacy's clientele* |  |  |  |  |
| University/colleges | 6 (30%) | 7 (41%) | 5 (28%) | 10 (50%) |
| Factory | 2 (10%) | 2 (12%) | 1 (6%) | 2 (10%) |
| Market | 15 (75%) | 14 (82%) | 16 (89%) | 17 (85%) |
| Bars/night clubs | 13 (65%) | 11 (65%) | 14 (78%) | 18 (90%) |
| Transit center | 6 (30%) | 7 (41%) | 11 (61%) | 9 (45%) |
| Central Business District | 7 (35%) | 3 (18%) | 3 (17%) | 3 (15%) |
| Walking distance between pharmacy and nearest public health facility |  |  |  |  |
| <15 minutes | 12 (60%) | 12 (70%) | 11 (61%) | 16 (80%) |
| 15-30 minutes | 6 (30%) | 2 (12%) | 3 (17%) | 2 (10%) |
| 30-45 minutes | 2 (10%) | 1 (6%) | 3 (17%) | 1 (5%) |
| >45 minutes | 0 (0%) | 2 (12%) | 1 (6%) | 1 (5%) |
| Age categories of clients served* |  |  |  |  |
| 16-24 years | 6 (30%) | 8 (47%) | 4 (22%) | 8 (40%) |
| 25-45 years | 19 (95%) | 15 (88%) | 17 (94%) | 19 (95%) |
| >45 years | 5 (25%) | 6 (35%) | 5 (28%) | 5 (25%) |
| Primary income bracket served |  |  |  |  |
| Low income (<24,000 KES/month) | 5 (25%) | 7 (41%) | 5 (28%) | 7 (35%) |
| Middle/high income (≥24,000 KES/month) | 15 (75%) | 10 (59%) | 13 (72%) | 13 (65%) |
| Key populations served* |  |  |  |  |
| Adolescent girls & young women (15-24 years) | 16 (80%) | 15 (88%) | 15 (83%) | 18 (90%) |
| Female sex workers | 7 (35%) | 11 (65%) | 6 (33%) | 9 (45%) |
| Clients of female sex workers | 6 (30%) | 8 (47%) | 5 (28%) | 8 (40%) |
| Men who have sex with men (MSM) | 3 (15%) | 2 (12%) | 3 (17%) | 2 (10%) |
| Migratory workers | 14 (70%) | 11 (65%) | 10 (56%) | 11 (55%) |
| Truck drivers | 8 (40%) | 8 (47%) | 7 (39%) | 4 (20%) |
| **Pharmacy staffing & structure** |  |  |  |  |
| Pharmacy ownership |  |  |  |  |
| Sole proprietor (1 single owner) | 16 (80%) | 14 (82%) | 17 (94%) | 13 (65%) |
| Partnership (2 or more owners) | 3 (15%) | 2 (12%) | 0 (0%) | 6 (30%) |
| Retail chain | 1 (5%) | 0 (0%) | 1 (6%) | 1 (5%) |
| Provider trained |  |  |  |  |
| Owner | 9 (45%) | 10 (59%) | 11 (61%) | 10 (50%) |
| Employee | 3 (15%) | 2 (12%) | 4 (22%) | 3 (15%) |
| Both owner and employee | 8 (40%) | 5 (29%) | 3 (17%) | 6 (30%) |
| Number of permanent staff | 1 (1-3) | 2 (1-2) | 2 (1-2) | 2 (1-2) |
| Pharmacy area* |  |  |  |  |
| Waiting area | 15 (75%) | 12 (71%) | 11 (61%) | 14 (70%) |
| Private exam room | 11 (55%) | 12 (71%) | 9 (50%) | 18 (90%) |
| Semi-private exam room | 9 (45%) | 6 (35%) | 9 (50%) | 2 (10%) |
| Other staff at the pharmacy* |  |  |  |  |
| Nurse(s) | 2 (10%) | 4 (24%) | 1 (6%) | 2 (10%) |
| *Other (e.g., clerk, courier)* | 7 (35%) | 12 (71%) | 5 (28%) | 9 (45%) |
| **Pharmacy services** |  |  |  |  |
| Prescriptions filled per week, on average |  |  |  |  |
| *0-499 prescriptions* | 15 (75%) | 11 (65%) | 17 (94%) | 15 (75%) |
| *500-999 prescriptions* | 5 (25%) | 5 (29%) | 1 (6%) | 4 (20%) |
| *>1000 prescriptions* | 0 (0%) | 1 (6%) | 0 (0%) | 1 (5%) |
| HIV prevention services offered prior to study launch* |  |  |  |  |
| *Condoms* | 20 (100%) | 17 (100%) | 18 (100%) | 20 (100%) |
| *HIV prevention counselling* | 14 (70%) | 12 (71%) | 15 (83%) | 15 (75%) |
| *HIV self-testing* | 18 (90%) | 16 (94%) | 16 (89%) | 19 (95%) |
| *HIV rapid diagnostic testing* | 4 (20%) | 3 (18%) | 5 (28%) | 5 (25%) |
| *PEP/PrEP/ART* | 0 (0%) | 0 (0%) | 0 (0%) | 0 (0%) |
| **Pharmacy system** |  |  |  |  |
| Inventory tracking system (any)* | 20 (100%) | 16 (94%) | 18 (100%) | 20 (100%) |
| *Paper records* | 10 (50%) | 7 (41%) | 6 (33%) | 9 (45%) |
| *Electronic records* | 12 (60%) | 11 (65%) | 14 (78%) | 14 (70%) |
| Refill reminders (any)* | 10 (50%) | 8 (47%) | 4 (22%) | 10 (50%) |
| *Phone call reminders* | 7 (35%) | 6 (35%) | 3 (17%) | 10 (50%) |
| *SMS/text reminders* | 6 (30%) | 3 (18%) | 3 (17%) | 9 (45%) |
| *WhatsApp message reminders* | 2 (10%) | 2 (12%) | 2 (11%) | 2 (10%) |

**Abbreviations:** Antiretroviral therapy (ART); interquartile range (IQR); men who have sex with men (MSM); post-exposure prophylaxis (PEP); pre-exposure prophylaxis (PrEP); short message service (SMS).
<sup>1</sup>All data reported in baseline pharmacy surveys
\* “Select all that apply” question.

### Participants

From June 26, 2023, to April 15, 2025, we enrolled 5,808 participants: 843 in the client-sustained, 2,254 in the implementor-sustained, 2,278 in the counselor-supported, and 433 in the clinic referral arms. At 60 days post-enrollment, 70% (n=4,061) of participants completed follow-up surveys (range 69-74% across arms; **Supplementary Figure 3**).

Participant characteristics are summarized in **Table 2**. Over half were male (54%, n=3,149) and unmarried (59%, n=3,401), with a median age of 27 (IQR 23, 34). Roughly two-thirds reported casual sexual partners (68%, n=3,924); few identified as a key population member (1.8%, n=102). The most common behaviors associated with HIV risk in the past six months were condomless sex (65%, n=3,785), sex with partners of unknown HIV status (53%, n=3,050), and multiple sexual partners (44%, n=2,546); condomless sex was also the most common potential HIV exposure in the past 72 hours (61%, n=3,531). Nearly all participants reported no prior PrEP use (96%, n=5,512) and almost two thirds were PEP candidates (with a potential HIV exposure in the past 72 hours; 64%, n=3,718).

**Table 2.** Characteristics of enrolled pharmacy clients, by study arm.

| Characteristics | 1: Client-sustained,<br>n=843 | 2: Implementor-sustained,<br>n=2,254 | 3: Counselor-supported,<br>n=2,278 | 4: Clinic referral (Control),<br>n=433 |
| --- | --- | --- | --- | --- |
| **Demographics** |  |  |  |  |
| Age <25 years old | 240/843 (28%) | 779/2,254 (35%) | 869/2,278 (38%) | 157/433 (36%) |
| Median age (IQR) | 29 (24–36) | 27 (23–34) | 27 (23–33) | 27 (23–33) |
| Male | 485/843 (58%) | 1,187/2,254 (53%) | 1,191/2,278 (52%) | 286/433 (66%) |
| Unmarried | 525/839 (63%) | 1,257/2,228 (56%) | 1,362/2,269 (60%) | 257/432 (59%) |
| Attended school | 187/839 (22%) | 359/2,228 (16%) | 475/2,268 (21%) | 129/432 (30%) |
| Median years attended school (IQR)<sup>1</sup> | 12 (12–15) | 13 (12–16) | 12 (10–15) | 12 (8–14) |
| Relationship status |  |  |  |  |
| Primary & casual partners | 217/839 (26%) | 707/2,228 (32%) | 837/2,269 (37%) | 186/432 (43%) |
| Casual only | 197/839 (23%) | 1,016/2,228 (46%) | 659/2,269 (29%) | 105/432 (24%) |
| Primary only | 253/839 (30%) | 416/2,228 (19%) | 688/2,269 (30%) | 114/432 (26%) |
| No partners | 172/839 (21%) | 89/2,228 (4.0%) | 85/2,269 (3.7%) | 27/432 (6.3%) |
| Population type |  |  |  |  |
| General population | 823/843 (98%) | 2,181/2,250 (97%) | 2,201/2,278 (97%) | 420/433 (97%) |
| Key population<sup>2</sup> | 1/843 (0.1%) | 41/2,250 (1.8%) | 54/2,278 (2.4%) | 6/433 (1.4%) |
| Serodiscordant couple | 19/843 (2.3%) | 28/2,250 (1.2%) | 23/2,278 (1.0%) | 7/433 (1.6%) |
| **Behaviors associated w/ HIV risk** |  |  |  |  |
| Past 6 months (PrEP eligibility) |  |  |  |  |
| Sex partner living with unmanaged<sup>3</sup> HIV (confirmed) | 41/843 (4.9%) | 75/2,251 (3.3%) | 156/2,278 (6.8%) | 23/433 (5.3%) |
| Sex partner at HIV risk (status unknown) | 316/843 (37%) | 1,070/2,251 (48%) | 1,466/2,278 (64%) | 198/433 (46%) |
| Multiple sex partners | 296/843 (35%) | 864/2,251 (38%) | 1,205/2,278 (53%) | 181/433 (42%) |
| Ongoing intimate partner violence | 38/843 (4.5%) | 47/2,251 (2.1%) | 102/2,278 (4.5%) | 6/433 (1.4%) |
| Transactional sex | 66/843 (7.8%) | 134/2,251 (6.0%) | 227/2,278 (10.0%) | 61/433 (14%) |
| Recent STI diagnosis/treatment | 54/843 (6.4%) | 56/2,251 (2.5%) | 120/2,278 (5.3%) | 20/433 (4.6%) |
| Recurrent PEP use | 0/843 (0%) | 0/2,251 (0%) | 0/2,278 (0%) | 0/433 (0%) |
| Sex with drugs/alcohol (recurrent) | 129/843 (15%) | 167/2,251 (7.4%) | 386/2,278 (17%) | 55/433 (13%) |
| Condomless sex | 466/843 (55%) | 1437/2,251 (64%) | 1606/2,278 (71%) | 276/433 (64%) |
| Injection drug use with shared needles | 9/843 (1.1%) | 25/2,251 (1.1%) | 19/2,278 (0.8%) | 8/433 (1.8%) |
| Past 72 hours (PEP eligibility) |  |  |  |  |
| Condomless sex | 658/843 (78%) | 1,299/2,251 (58%) | 1,266/2,278 (56%) | 308/433 (71%) |
| Shared needles | 21/843 (2.5%) | 40/2,251 (1.8%) | 39/2,278 (1.7%) | 6/433 (1.4%) |
| Sexual assault | 19/843 (2.3%) | 23/2,251 (1.0%) | 25/2,278 (1.1%) | 7/433 (1.6%) |
| Other high risk | 16/843 (1.9%) | 34/2,251 (1.5%) | 48/2,278 (2.1%) | 6/433 (1.4%) |
| **Perceived HIV risk (self-assessment)** |  |  |  |  |
| Currently at HIV risk | 414/843 (49%) | 1,272/2,251 (57%) | 1,351/2,278 (59%) | 224/433 (52%) |
| Future HIV risk (next month): high | 72/843 (8.5%) | 571/2,251 (25%) | 526/2,277 (23%) | 132/433 (30%) |
| **Prior PrEP use** |  |  |  |  |
| No prior PrEP use | 806/839 (96%) | 2,143/2,228 (96%) | 2,140/2,269 (94%) | 423/432 (98%) |
| **PrEP or PEP candidacy<sup>4</sup>** |  |  |  |  |
| PrEP candidate | 2,075/5,793 (36%) | 157/842 (19%) | 862/2,249 (38%) | 908/2,270 (40%) |
| PEP candidate | 3,718/5,793 (64%) | 685/842 (81%) | 1,387/2,249 (62%) | 1,362/2,270 (60%) |

**Abbreviations:** Interquartile range (IQR); Pre-exposure prophylaxis (PrEP); post-exposure prophylaxis (PEP); sexually transmitted infection (STI).
<sup>1</sup> Number of observations missing in each arm: 1: Client-sustained (21); 2: Implementor-sustained (73); 3: Counselor-supported (39); 4: Clinic-referral (59).
<sup>2</sup> Key populations are groups that are particularly vulnerable to HIV, and include: men who have sex with men, sex workers, people who inject drugs, and transgender people.
<sup>3</sup> Not taking antiretrovirals to keep viral load undetectable and non-transmissible.
<sup>4</sup> 15 clients were neither PrEP or PEP candidates.

### Primary PrEP outcomes

At 60 days post-enrollment, 2,214 participants self-reported PrEP initiation and 803 one-month PrEP continuation (36% of all initiators; **Table 3**). Mean standardized annual pharmacy PrEP initiation rates were 4.6 (standard deviation [SD] 7.8) in the client-sustained, 17 (SD 69) in the implementor-sustained, 26 (SD 33) in the counselor-supported, and 2.7 (SD 6.2) in the clinic referral arms. Compared to clinic referral, PrEP initiation rates were significantly higher in the counselor-supported arm (RR=6.5, 95% CI [2.6, 16], p<0.001) and higher, though not significantly so at the Bonferroni threshold, in the implementor-sustained arm (RR=4.3, 95% CI [1.1, 16], p=0.035; **Figure 3a**). The client-sustained arm did not significantly differ from clinic referral (RR=1.4, 95% CI [0.52, 3.9], p=0.49). Mean standardized annual pharmacy PrEP continuation rates within 60 days of enrollment were 0.98 (SD 1.6), 7.1 (SD 31), 8.6 (SD 9.8) and 1.1 (SD 3.3) in the client-sustained, implementor-sustained, counselor-supported, and clinic referral arms, respectively. Only the counselor-supported arm demonstrated a significantly higher one-month PrEP continuation rate than clinic referral (RR=5.1, 95% CI [1.4, 19], p=0.016); the implementor-sustained arm trended toward a higher PrEP continuation rate, with a large point estimate (RR=4.5, 95% CI [0.77, 26], p=0.093), but wide confidence intervals included the null.

**Figure 3a.**
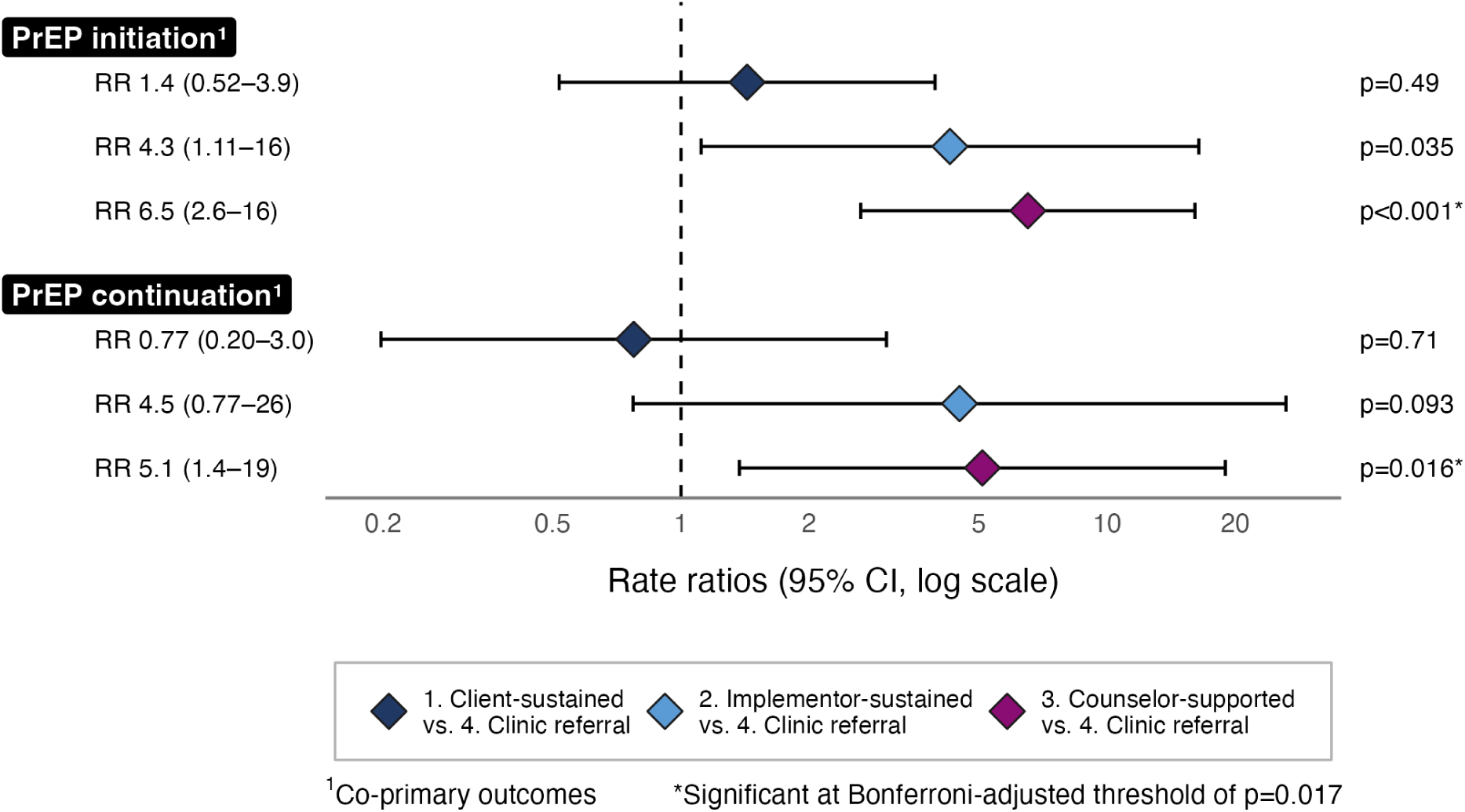
Primary PrEP outcome rate ratios at 60 days post-enrollment.

**Table 3.** Description of pharmacy volume and study outcomes 60 days from enrollment, by study arm.

| Outcomes<sup>1</sup> |  | 1: Client-sustained | 2: Implementor-sustained | 3: Counselor-supported | 4: Clinic referral (control) |
| --- | --- | --- | --- | --- | --- |
| **PrEP outcomes** |  |  |  |  |  |
| Initiation (*primary*) | n | 169 | 949 | 978 | 118 |
|  | Mean annual pharmacy rate (SD)<sup>2</sup> | 4.6 (7.8) | 17 (69) | 26 (33) | 2.7 (6.2) |
| Continuation (*primary*) | n | 36 | 400 | 319 | 48 |
|  | Mean annual pharmacy rate (SD)<sup>2</sup> | 0.98 (1.6) | 7.1 (31) | 8.6 (9.8) | 1.1 (3.3) |
| **PEP outcomes** |  |  |  |  |  |
| Initiation | n | 684 | 1,364 | 1,367 | 250 |
|  | Mean annual pharmacy rate (SD)<sup>2</sup> | 19 (19) | 24 (41) | 37 (28) | 5.8 (7.4) |
| Recurrent PEP use | n | 4 | 17 | 25 | 2 |
|  | Mean annual pharmacy rate (SD)<sup>2</sup> | 0.11 (0.40) | 0.30 (0.95) | 0.68 (1.2) | 0.05 (0.18) |
| PEP-to-PrEP transition | n | 16 | 110 | 85 | 4 |
|  | Mean annual pharmacy rate (SD)<sup>2</sup> | 0.43 (0.86) | 2.0 (7.9) | 2.3 (3.2) | 0.09 (0.25) |
| **PrEP/PEP outcomes** |  |  |  |  |  |
| Initiation | n | 831 | 2,218 | 2,249 | 341 |
|  | Mean annual pharmacy rate (SD)<sup>2</sup> | 23 (22) | 39 (101) | 61 (45) | 7.9 (12) |
| Continuation or recurrent use | n | 60 | 526 | 445 | 50 |
|  | Mean annual pharmacy rate (SD)<sup>2</sup> | 1.6 (2.9) | 9.4 (41) | 12 (12) | 1.2 (2.9) |

**Abbreviations:** Post-exposure prophylaxis (PEP); pre-exposure prophylaxis (PrEP); standard deviation (SD).
<sup>1</sup>Missing survey data were imputed using observed outcome rates in the same pharmacy (a post-hoc analytic decision based on evidence that our assumption that missing=failure was invalid). The results corresponding to the pre-specified analysis in which missing outcomes were imputed as failures are presented in Supplementary Table 3.
<sup>2</sup>Mean annual pharmacy rates were weighted by client volume and standardized to the median pharmacy volume in the clinic referral arm (2,030 clients/month).

Sensitivity analyses incorporating implementation pauses and permutation tests were consistent with the primary findings: only the counselor-supported arm demonstrated significantly higher PrEP initiation and continuation rates than clinic referral, and the implementor-sustained arm trended toward higher rates (**Supplementary Table 1**). Among PrEP initiators, the proportion continuing PrEP did not significantly differ between any direct delivery arm and clinic referral. Subgroup analyses by region and urbanicity revealed no significant differences in arm comparisons, though PrEP initiation and continuation rates were notably higher in Western than Central Kenya, albeit with sample sizes (**Supplementary Table 2**). Primary and secondary findings (described below) were largely unchanged when applying the original assumption that missing client survey data represented failures (**Supplementary Table 3**).

### Secondary PEP outcomes

At 60 days post-enrollment, 3,665 participants self-reported PEP initiation, representing 65% of all PrEP/PEP initiations (n=5,639); recurrent PEP use and PEP-to-PrEP transition were self-reported by 48 (1.3%) and 215 (5.9%) of PEP initiators, respectively (**Table 3**). Mean standardized annual pharmacy PEP initiation rates were 19 (SD 19), 24 (SD 41), 37 (SD 28), and 5.8 (SD 7.4) in the client-sustained, implementor-sustained, counselor-supported, and clinic referral arms, respectively. All direct delivery arms demonstrated significantly higher PEP initiation rates than clinic referral (client-sustained: RR=3.5, 95% CI [1.4, 8.8], p=0.010; implementor-sustained: RR=4.0, 95% CI [1.4, 11], p=0.008; counselor-supported: RR=6.0, 95% CI [2.4, 15], p<0.001; **Figure 3b**). Recurrent PEP use and PEP-to-PrEP transition rates were low across all study arms at 60 days. Only the counselor-supported arm demonstrated significantly higher recurrent PEP use than clinic referral (RR=14, 95% CI [2.2, 97], p=0.007); both the implementor-sustained and counselor-supported arms demonstrated significantly higher PEP-to-PrEP transition rates (implementor-sustained: RR=15, 95% CI [3.0, 79], p=0.001; counselor-supported: RR=17, 95% CI [4.5, 67], p<0.001). Wide confidence intervals for these outcomes reflect their low prevalence.

**Figure 3b.**
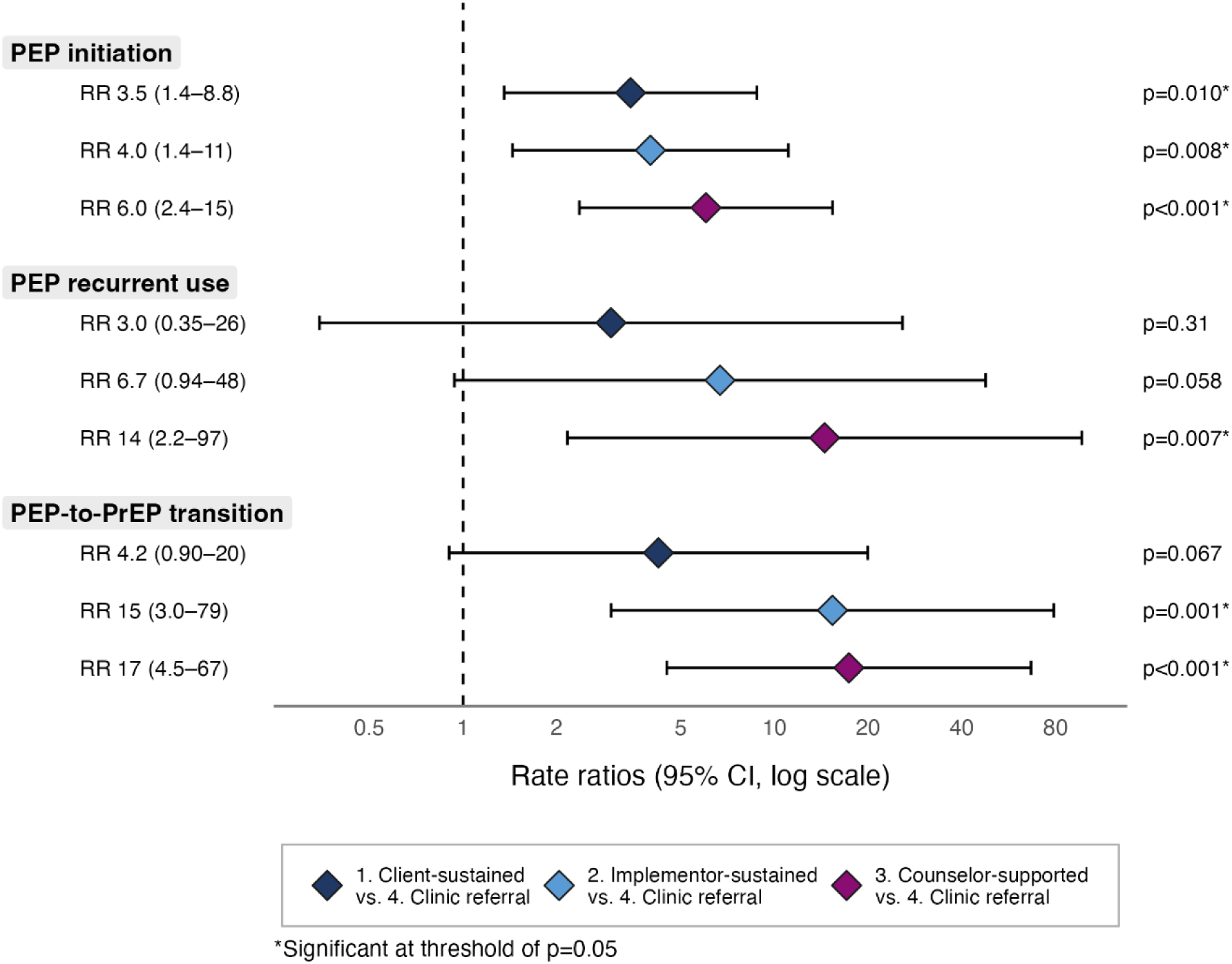
Secondary PEP outcome rate ratios at 60 days post-enrollment.

### Secondary PrEP/PEP outcomes

At 60 days post-enrollment, 5,639 participants reported PrEP/PEP initiation and 1,081 one-month PrEP/PEP continuation (19% of all initiators; **Table 3**). Mean standardized annual pharmacy PrEP/PEP initiation rates were 23 (SD 22), 39 (SD 101), 61 (SD 45), and 7.9 (SD 12) in the client-sustained, implementor-sustained, counselor-supported, and clinic referral arms, respectively; mean standardized annual continuation rates were 1.6 (SD 2.9), 9.4 (SD 41), 12 (SD 12), and 1.2 (SD 2.9), respectively. Compared to clinic referral, PrEP/PEP initiation rates were significantly higher across all direct delivery arms (client-sustained: RR=2.8, 95% CI [1.24, 6.4], p=0.014; implementor-sustained: RR=4.3, 95% CI [1.56, 12], p=0.005; counselor-supported: RR=6.5, 95% CI [3.0, 14], p<0.001; **Figure 3c**); significantly higher continuation rates were observed only in the counselor-supported arm (RR=7.0, 95% CI [2.4, 20], p<0.001).

**Figure 3c.**
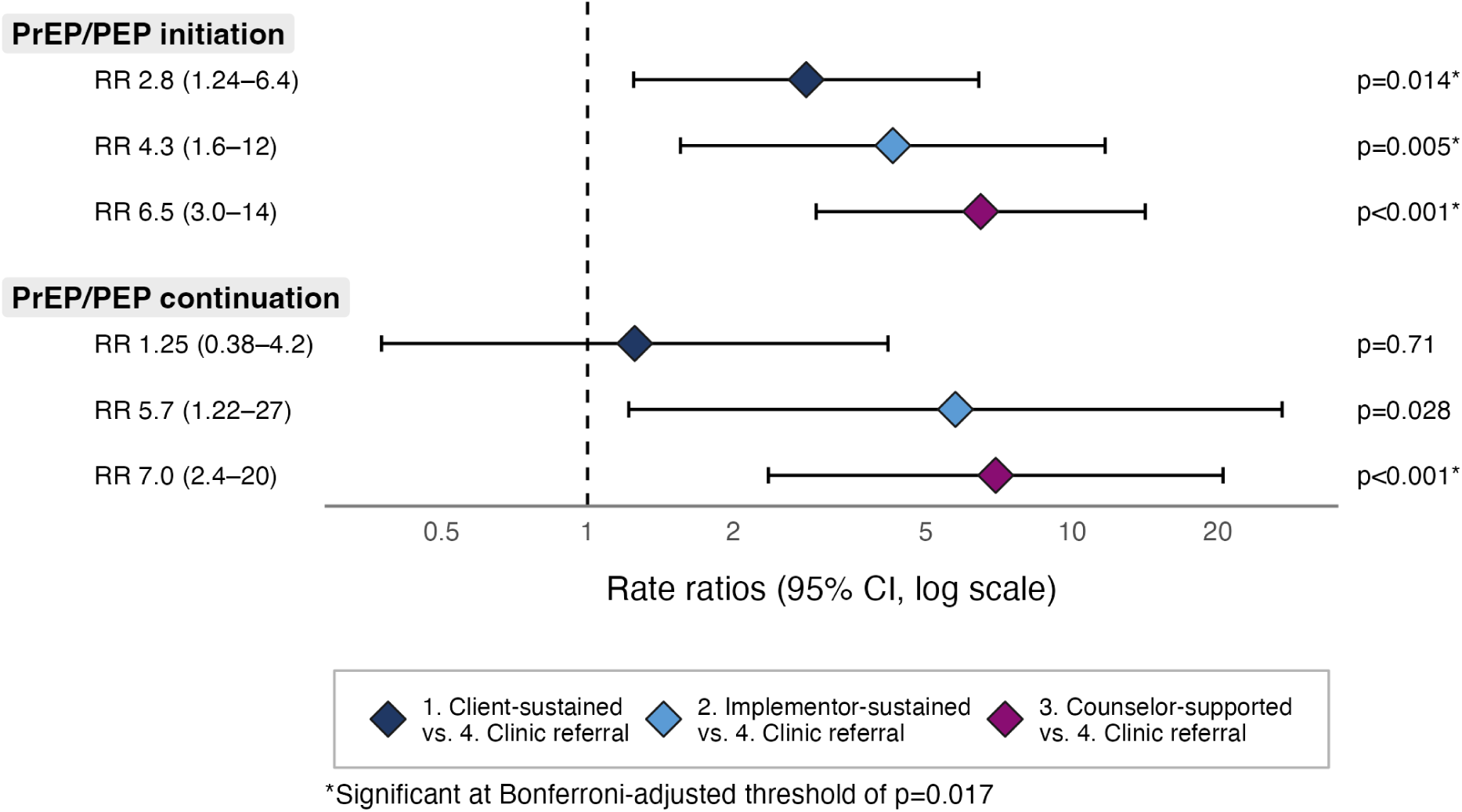
Secondary PrEP/PEP outcome rate ratios at 60 days post-enrollment.

### Pharmacy-specific performance

Pharmacy-specific performance varied considerably across and within arms (**Figure 4**). Despite training and routine technical assistance, a subset of pharmacies showed low intervention engagement, initiating or continuing few or no clients on PrEP throughout implementation. Conversely, a subset of high-performing pharmacies—particularly in the implementor-sustained arm—were outliers, likely driving the wide confidence intervals observed for that arm. PEP initiation was more consistently achieved across pharmacies, though rates still spanned a wide range within each arm.

**Figure 4.**
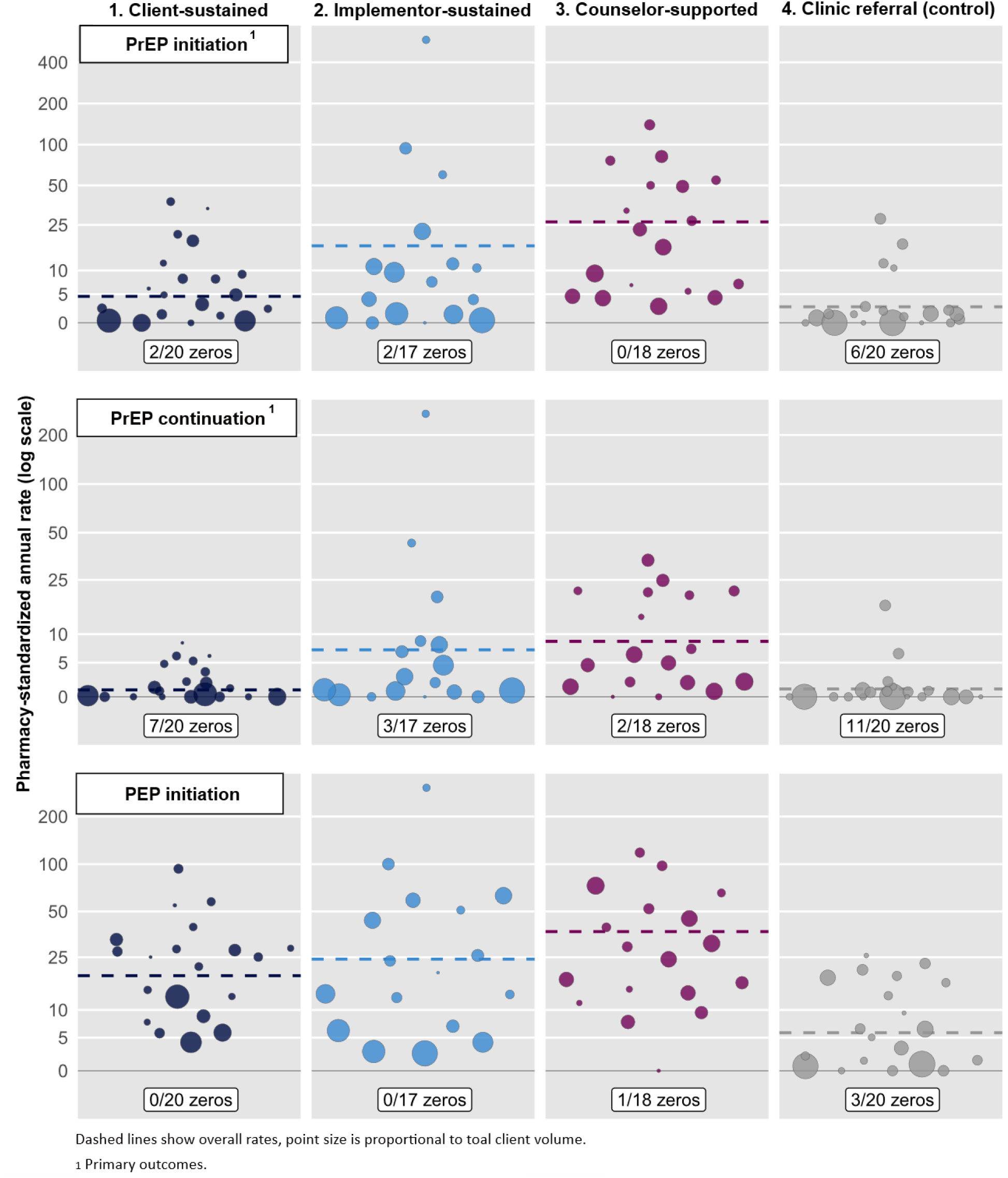
Pharmacy-specific performance: PrEP initiation, PrEP continuation, and PEP initiation rates at 60 days post-enrollment^1^.

### HIV seroconversions and adverse events

Over the study duration, one HIV seroconversion (client-sustained arm), one social harm (relationship conflict; counselor-supported arm), and two provider needlestick injuries (counselor-supported and client-sustained arms) were reported. All participants were linked to follow-up care except the participant who reported the social harm, who requested no further contact.

## DISCUSSION

In this first randomized trial testing different models of private pharmacy provider-led initiation and management of PrEP and PEP, direct pharmacy-based delivery engaged more clients than clinic referral, particularly when services were fully subsidized and time-intensive delivery steps task-shifted to HTS counselors. Our findings also suggest greater client price sensitivity to PrEP versus PEP: user fees had less of an impact on PEP initiation—a time-sensitive post-exposure intervention—than on PrEP initiation, which requires recognizing and acting on anticipated HIV risk before an exposure occurs. Notably, PEP demand was high, with most clients initiating PEP over PrEP, underscoring private pharmacies’ capacity to reach individuals with urgent post-exposure needs. Finally, heterogeneity in pharmacy-level performance suggests pharmacy-based PrEP/PEP delivery may not suit all settings, pointing to the need to refine pharmacy eligibility criteria and prioritize pharmacies best positioned to sustain delivery.

Findings from trials of other pharmacy-delivered preventive SRH products align with our findings and offer insights into strategies to improve PrEP and PEP outcomes.^30,31^ In Kenya, a cRCT of 137 pharmacies found that client subsidies and provider incentives significantly increased uptake of a long-acting injectable contraceptive while reducing uptake of short-acting methods.^30^ A cRCT of 29 pharmacies in the United Kingdom (UK) found that offering an advance supply of daily oral progestogen contraception alongside emergency contraception increased uptake of effective ongoing contraception.^31^ The Kenyan cRCT findings align with ours: fully subsidized services and task-shifting to HTS counselors—which reduced provider workload and, in effect, incentivized delivery—drove the largest gains in PrEP and PEP initiation. The UK cRCT further suggests that advance PrEP dispensing at PEP initiation might improve the low PEP-to-PrEP transition rates observed in our study.

As external funding for HIV programs contracts, governments face growing pressure to sustain HIV responses and are increasingly considering private-sector delivery.^32,33^ Our study’s client-sustained delivery arm aligns with the Kenya MOH’s vision for private-sector partnership:^6^ a mixed-financing model in which government supplies free PrEP/PEP drugs from national stock and clients pay a user fee for counseling, dispensing, and follow-up services. Our findings, however, indicate that eliminating this fee yields greater PrEP and PEP initiation and continuation. The financial case for a full subsidy model is also compelling: every HIV infection prevented avoids a lifetime of antiretroviral treatment costs typically borne by government. Complimentary costing research suggests that while partially subsidized delivery has lower total costs than fully subsidized delivery, it reaches fewer clients, yielding higher per-visit costs.^14^ Where a full subsidy approach is infeasible, targeted strategies—such as vouchers^34^ or coverage by the national health insurance agency^35^—could eliminate user fees for populations most likely to benefit (e.g., young people <25 years).

Our findings also highlight the benefits of task-shifting the time-intensive PrEP/PEP delivery steps—HIV testing and counseling—away from overburdened pharmacy providers to a dedicated worker. Complementary qualitative research suggests counselor-supported clients also received more comprehensive counseling, with fewer demonstrating PrEP/PEP misconceptions.^36^ We selected HTS counselors for their lower cost relative to nurses or clinicians^37,38^ and historical availability across Africa through support from the U.S. President’s Emergency Plan for AIDS Relief (PEPFAR).^39,40^ However, complementary costing research suggests that stationing a full-time counselor at each pharmacy is costly—particularly at lower-volume sites—limiting scalability,^15^ and ongoing PEPFAR cuts further threaten this workforce’s availability.^41^ Lower-cost task-shifting strategies are therefore needed, especially as the growing number of PrEP products available in community settings increases counseling complexity. Promising approaches include HIV self-testing (shifting testing to clients),^42^ telemedicine (enabling one remote counselor or nurse to support multiple pharmacies),^43^ and AI-powered tools (enhancing between-visit adherence and counseling support).^44,45^ Several of these strategies are currently under evaluation (NCT07561658).^43^

These findings emphasize that pharmacies can reach PEP clients with time-sensitive needs, consistent with family planning literature showing higher emergency contraception dispensing at private pharmacies than public clinics.^46^ With extended hours and convenient locations, pharmacies are well-positioned to ensure clients access PEP within 72 hours—ideally within 24 hours—of a potential HIV exposure. These findings also underscore PEP’s important yet underutilized role in HIV prevention programming.^47^ For individuals with infrequent or unpredictable sexual encounters, for whom anticipating HIV risk is difficult, time-limited PEP may be preferable to continuous PrEP. Modeling studies show community-based PEP provision may be more cost-effective than PrEP, given its alignment with HIV risk exposure and short use duration.^48^ HIV programs should therefore prioritize rapid PEP access and develop new strategies (e.g., advance drug dispensing^49,50^) to support clients with ongoing risk in transitioning to PrEP. Finally, as long-acting antiretrovirals—including monthly oral formulations^51^—advance through clinical development, pharmacies could serve as important venues for both effectiveness research and eventual scale-up of novel PEP regimens.^52^

We observed substantial variation in pharmacy-specific PrEP/PEP performance, suggesting heterogeneity in context, clientele, and providers can influence implementation. Pharmacies varied in setting—near universities and bars versus business districts or transit centers—influencing client reach and retention. Additionally, since most pharmacies operated with a single pharmacy provider per shift, pharmacy-level outcomes were likely highly influenced by individual provider engagement. This may produce greater variability than in clinic-based PrEP programs, where success often hinges on a “PrEP champion”—a provider who dedicates themself to promoting and driving implementation.^53^ Qualitative research is ongoing to further explore drivers of pharmacy performance. Our findings suggest that this intervention can yield population-level benefits even when some pharmacies perform poorly (i.e., initiate and manage few clients); implementation success and public health impact instead likely depend on selecting and enabling pharmacies with adequate reach and staffing, and on identifying a financing model that motivates providers while remaining affordable to clients.

This trial has several strengths and limitations. Strengths include implementation across a large sample of real-world pharmacies in diverse geographic settings, increasing generalizability; offering both PrEP and PEP to address clients’ varied HIV prevention needs; testing three distinct pharmacy-based delivery models to inform scale-up strategies; and implementing services in a manner that closely mirrors real-world delivery—integrating pharmacies into Kenya’s PrEP/PEP supply chain, training providers in collaboration with MOH officials, and positioning RAs offsite to avoid biasing implementation. Limitations include reliance on self-reported PrEP and PEP outcomes, necessary given the clinic referral arm; inability to determine the number of clients at each pharmacy eligible for PrEP or PEP—a challenge inherent to HIV prevention research—necessitating reliance on overall pharmacy client volume as a proxy; pharmacy client volume not being measured a priori or accounted for in randomization, requiring post-hoc measurement and model inclusion; relatively few clusters per arm, typical of cRCTs; insufficient power to compare the direct delivery arms to one another; 25% (15/60) pharmacy turnover, reducing implementation time; and a short follow-up period—appropriate for assessing initiation and limiting recall bias, but less suitable for assessing longer-term PrEP engagement and recurrent PEP use (270-day outcomes will be reported separately).

Pharmacy-based PrEP/PEP delivery is nascent but growing;^54-56^ this study fills critical evidence gaps by demonstrating that direct pharmacy-based PrEP and PEP delivery outperforms clinic referral with minimal training and support, and that eliminating client user fees and task-shifting time-intensive delivery steps improves outcomes. Our findings further establish pharmacies’ capacity to reach clients with time-sensitive post-exposure needs and reveal substantial heterogeneity in pharmacy-level performance, underscoring the need to refine eligibility criteria and prioritize pharmacies best positioned to sustain delivery. Future research should explore strategies to optimize pharmacy-based PrEP/PEP services—including targeted subsidies,^57^ telemedicine-based task-shifting,^10^ and advance PrEP dispensing to support PEP-to-PrEP transition^49,50^—as well as pharmacies’ potential as platforms for long-acting HIV prevention products (e.g., semiannual injections), HIV treatment, and complementary SRH services (e.g., STI testing). As donor funding for HIV programs contracts,^41^ innovative and low-cost delivery strategies that expand the reach of prevention services are more critical than ever. Private pharmacies should be prioritized for inclusion in HIV prevention programs.

## DECLARATIONS

### Contributors

KFO, KN, EAB, KC and DW wrote the proposal and secured the funding for this trial. KFO, SR, KN, and EAB drafted the study protocol. RM, KH, DW, TK, VO obtained ethical approvals. SR, KFO developed the study forms. TK, CK, VO, PAO, LJ, PO, MOA, EAB, DW, KN oversaw all implementation and collection of study data. AM, KFO, KY, PB developed the statistical analysis plan for this trial. PB, KY, EG, MA, KT, TS managed, accessed, verified, and analyzed the data. KFO, KH, SR, RM drafted the manuscript, which was reviewed and edited by all authors. All authors had full access to all the data in the study and had final responsibility for the decision to submit for publication.

### Data sharing

Deidentified participant and pharmacy data supporting the conclusions of this article are available in the Harvard Dataverse repository, [*unique persistent identifier and hyperlink to dataset in https:// format—will update following review*]^58^ immediately following publication.

## Acknowledgements

We thank all trial participants—including pharmacy clients and providers—for their time and contributions to this research. We are grateful to the technical officers at Jhpiego Kenya (June Mwangi, Doreen Muriithi, Sharon Chebet, Gordon Okello) and the research staff at the Kenyan Medical Research Institute in Thika and Kisumu for their dedication to the trial. We thank colleagues at the Kenya Ministry of Health and on the County Health Management Teams for their support and assistance selecting study pharmacies and securing commodities for implementation. We also thank the team at Maisha Meds (Jess Vernon, Elizabeth Kathure,

Wintana Balai) for programming and prescribing checklist and monthly commodity tracking reports. Finally, we thank members of our Data Safety and Monitoring Committee members (Deborah Donnell, John Kinuthia, Eric Sedah, Peter Mugo) for their guidance in navigating implementation challenges and ensuring successful trial completion.

