## Supplementary material for "Effectiveness of pharmacy-based HIV pre- and post-exposure prophylaxis delivery: a cluster-randomized trial in Kenya": Supp tables and figures

### Figure titles:

**Supplementary Figure 1. Estimated monthly client volume and months of implementation by study pharmacies and arm.**

**Supplementary Figure 2. Screening, enrollment, and completion of 60-day surveys for pharmacy clients, by arm**

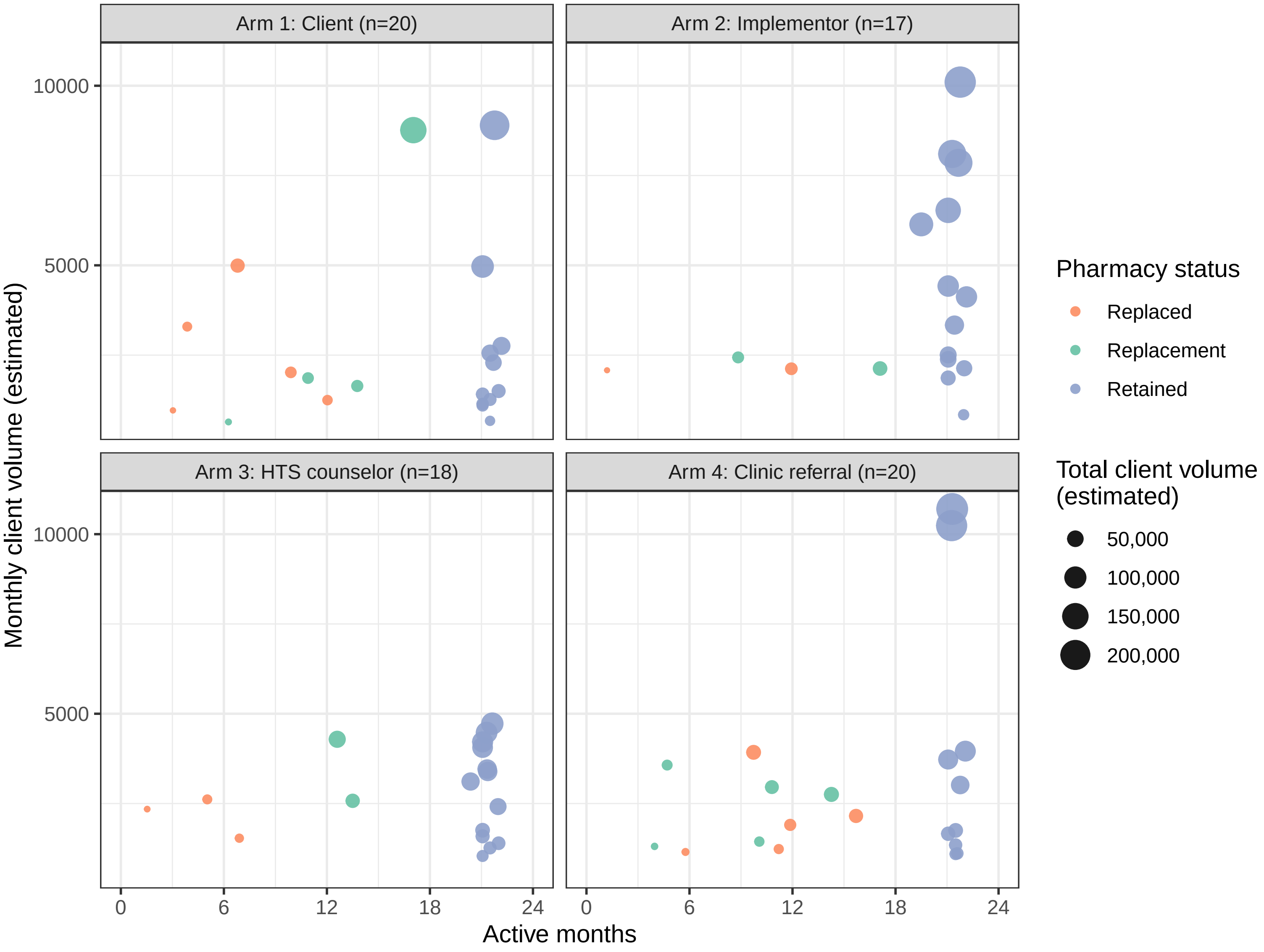

Arm 1: Client (n=20)
Arm 2: Implementor (n=17)
Arm 3: HTS counselor (n=18)
Arm 4: Clinic referral (n=20)
Monthly client volume (estimated)
10000
5000
0
6
12
18
24
Active months
Pharmacy status
Replaced
Replacement
Retained
Total client volume (estimated)
50,000
100,000
150,000
200,000

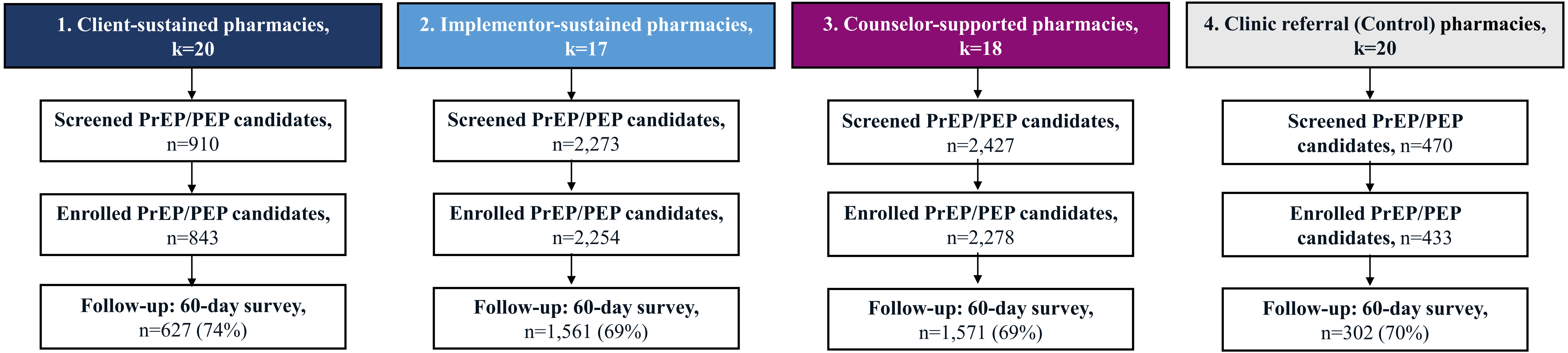


Abbreviations: Pre-exposure prophylaxis (PrEP); post-exposure prophylaxis (PEP).

### Supplementary Table 1: Pre-specified sensitivity analyses and select secondary analyses for primary study outcomes

| Outcomes | | Survey data | | | | Effect size estimates | | | | | |
|---|---|---|---|---|---|---|---|---|---|---|---|
| | | 1: Client-sustained | 2: Implementor-sustained | 3: Counselor-supported | 4: Clinic referral (Control) | 1: Client-sustained vs. 4: Clinic referral | | 2: Implementor-sustained vs. 4: Clinic referral | | 3: Counselor-supported vs. 4: Clinic referral | |
| **<u>Sensitivity 1</u>. Temporary pharmacy implementation pauses incorporated[3]** | | | | | | **RR (95% CI)[1]** | ***p*** | **RR (95% CI)[1]** | ***p*** | **RR (95% CI)[1]** | ***p*** |
| PrEP initiation* | n | 169 | 949 | 978 | 118 | | | | | | |
| | Mean annual rate[2] | 4.7 | 18 | 27 | 2.8 | 1.40 (0.51–3.9) | 0.51 | 4.3 (1.11–17) | 0.035 | 6.5 (2.6–16) | <0.001 |
| PrEP continuation* | n | 36 | 400 | 319 | 48 | | | | | | |
| | Mean annual rate[2] | 1.0 | 7.5 | 8.9 | 1.2 | 0.76 (0.19–3.0) | 0.69 | 4.5 (0.77–27) | 0.093 | 5.1 (1.36–19) | 0.016 |
| **<u>Sensitivity 2</u>. Non-parametric permutation testing[4]** | | | | | | | ***p*** | | ***p*** | | ***P*** |
| PrEP initiation* | | -- | -- | -- | -- | -- | 0.65 | -- | 0.031 | -- | 0.003 |
| PrEP continuation* | | -- | -- | -- | -- | -- | 0.83 | -- | 0.059 | -- | 0.019 |
| **<u>Sensitivity 3</u>. High-volume pharmacies excluded[5]** | | | | | | **RR (95% CI)[1]** | ***p*** | **RR (95% CI)[1]** | ***p*** | **RR (95% CI)[1]** | ***p*** |
| PrEP initiation* | n | 164 | 867 | 978 | 118 | | | | | | |
| | Mean annual rate[2] | 7.2 | 38 | 26 | 4.8 | 1.35 (0.41–4.5) | 0.62 | 9.1 (0.59–142) | 0.11 | 5.0 (1.58–16) | 0.007 |
| PrEP continuation* | n | 32 | 353 | 319 | 48 | | | | | | |
| | Mean annual rate[2] | 1.4 | 16 | 8.6 | 1.9 | 0.68 (0.12–3.7) | 0.65 | 15 (0.20–1070) | 0.22 | 3.8 (0.77–19) | 0.10 |
| **<u>Secondary</u>: PrEP continuation among PrEP initiators[6]** | | | | | | **ADP (95% CI)** | ***p*** | **ADP (95% CI)** | ***p*** | **ADP (95% CI)** | ***p*** |
| PrEP continuation* | n/N | 27/142 | 270/670 | 243/700 | 31/79 | | | | | | |
| | Proportion | 0.19 | 0.40 | 0.35 | 0.39 | -0.22 (-0.50–0.07) | 0.12 | -0.03 (-0.35–0.29) | 0.83 | -0.09 (-0.40–0.21) | 0.48 |

**Abbreviations:** Absolute difference in proportions (ADP); Confidence interval (CI); Interquartile range (IQR); post-exposure prophylaxis (PEP); pre-exposure prophylaxis (PrEP); rate ratios (RR).

[1]We estimated rate ratios using quasi-Poisson generalized linear models with a log link, modeling outcome counts with an offset for the total client volume and adjusting for county group. 95% CIs and p-values use HC3 robust standard errors. Missing survey data were imputed using observed outcome rates in the same pharmacy (a post-hoc analytic decision based on evidence that our assumption that missing=failure was invalid).

[2]Mean annual pharmacy rates were weighted by client volume and standardized to the median pharmacy volume in the clinic referral arm (2,030 clients/month).

[3]In this sensitivity analysis, we accounted for pharmacies' temporary pauses in implementation, excluding that time when calculating pharmacy-specific rates and client-volume offset in the quasi-Poisson models.

[4]In this sensitivity analysis, nonparametric permutation tests were used to assess the robustness of the findings from the original analysis with fewer distributional assumptions.

[5]In this sensitivity analysis, we excluded all high-volume pharmacies (monthly client volume > 5,000). This was a post-hoc analysis, added because arm 3 did not contain any high-volume pharmacies.

[6]In this secondary analysis we compared PrEP continuation rates among PrEP initiators, at the individual level. Because continuation is conditional on a post-randomization event (initiation) we adjusted for county group, age, sex, and HIV risk self-perception to address potential selection bias. We estimated risk differences using Gaussian generalized estimating equation (GEE) models with identity link and independence working correlation. 95% CIs and p-values use CR2 cluster-robust standard errors with Satterthwaite degrees of freedom to account for the small number of clusters.

### Supplementary Table 2: Pre-specified regional and urbanicity sub-group analyses for our primary study outcomes

| Outcomes | | Survey data | | | | Effect size estimates[1] | | | | | |
|---|---|---|---|---|---|---|---|---|---|---|---|
| | | 1: Client-sustained | 2: Implementor-sustained | 3: Counselor-supported | 4: Clinic referral | 1: Client-sustained vs. 4: Clinic referral | | 2: Implementor-sustained vs. 4: Clinic referral | | 3: Counselor-supported vs. 4: Clinic referral | |
| **PrEP initiation** | | | | | | **RR (95% CI)** | ***p***[3] | **RR (95% CI)** | ***p***[3] | **RR (95% CI)** | ***p***[3] |
| <u>Regional subgroups</u>: | | | | | | | *0.28* | | *0.95* | | *0.35* |
| Central Kenya, k=37 | n | 57 | 92 | 166 | 21 | | | | | | |
| | Mean annual rate[2] | 2.2 | 2.6 | 7.4 | 0.62 | 3.5 (0.34–36) | 0.45 | 4.2 (0.66–27) | 0.17 | 12 (2.5–58) | <0.001 |
| Western Kenya, k=38 | n | 112 | 857 | 812 | 97 | | | | | | |
| | Mean annual rate[2] | 11 | 41 | 55 | 10 | 1.03 (0.28–3.8) | 1.00 | 3.9 (0.59–26) | 0.21 | 5.3 (1.51–19) | 0.005 |
| <u>Urbanity subgroups</u>: | | | | | | | *0.62* | | *0.11* | | *0.089* |
| Urban, k=61 | n | 161 | 780 | 927 | 38 | | | | | | |
| | Mean annual rate[2] | 4.5 | 21 | 28 | 1.1 | 2.1 (0.43–9.8) | 0.55 | 11 (0.73–152) | 0.097 | 10 (2.7–38) | <0.001 |
| Rural/peri-urban, k=14 | n | 8 | 169 | 51 | 80 | | | | | | |
| | Mean annual rate[2] | 5.7 | 9.1 | 12 | 8.5 | 1.17 (0.18–7.8) | 0.98 | 1.16 (0.31–4.4) | 0.97 | 2.2 (0.41–12) | 0.55 |
| **PrEP continuation** | | | | | | **RR (95% CI)** | ***p***[3] | **RR (95% CI)** | ***p***[3] | **RR (95% CI)** | ***p***[3] |
| <u>Regional subgroups</u>: | | | | | | | *0.23* | | *0.64* | | *0.50* |
| Central Kenya, k=37 | n | 14 | 66 | 53 | 9 | | | | | | |
| | Mean annual rate[2] | 0.53 | 1.9 | 2.4 | 0.27 | 2.0 (0.27–15) | 0.73 | 7.0 (0.90–55) | 0.068 | 8.9 (1.54–51) | 0.009 |
| Western Kenya, k=38 | n | 22 | 334 | 266 | 39 | | | | | | |
| | Mean annual rate[2] | 2.1 | 16 | 18 | 4.2 | 0.50 (0.079–3.2) | 0.69 | 3.8 (0.35–42) | 0.41 | 4.3 (0.75–25) | 0.13 |
| <u>Urbanity subgroups</u>: | | | | | | | *--*[4] | | *0.067* | | *0.34* |
| Urban, k=61 | n | 35 | 347 | 294 | 10 | | | | | | |
| | Mean annual rate[2] | 0.99 | 9.2 | 9.0 | 0.30 | 2.2 (0.42–11) | 0.53 | 21 (0.65–673) | 0.10 | 14 (2.8–67) | <0.001 |
| Rural/peri-urban, k=14 | n | 1 | 53 | 25 | 38 | | | | | | |
| | Mean annual rate[2] | 0.71 | 2.9 | 5.7 | 4.0 | --[4] | --[4] | 0.67 (0.099–4.6) | 0.90 | 2.4 (0.046–123) | 0.89 |

**Abbreviations:** Confidence interval (CI); post-exposure prophylaxis (PEP); pre-exposure prophylaxis (PrEP); rate ratios (RR).

[1]We estimated rate ratios using quasi-Poisson generalized linear models with a log link, modeling outcome counts with an offset for the total client volume and adjusting for county group. 95% CIs and p-values use HC3 robust standard errors. Missing survey data were imputed using observed outcome rates in the same pharmacy (a post-hoc analytic decision based on evidence that our assumption that missing=failure was invalid).

[2]Mean annual pharmacy rates were weighted by client volume and standardized to the median pharmacy volume in the clinic referral arm (2,030 clients/month).

[3] Italicized p-values in subgroup headers are for arm × subgroup interaction terms.

[4] Numbers too small to estimate.

**Supplementary Table 3: Rate ratios for study outcomes resulting from the pre-specified analysis in which unobserved outcomes were imputed as failures.**

| **Outcomes** | | **Survey data** | | | | **Effect size estimates[1]** | | | | | |
|---|---|---|---|---|---|---|---|---|---|---|---|
| | | **1: Client-sustained** | **2: Implementor-sustained** | **3: Counselor-supported** | **4: Clinic referral** | **1: Client-sustained vs. 4: Clinic referral** | | **2: Implementor-sustained vs. 4: Clinic referral** | | **3: Counselor-supported vs. 4: Clinic referral** | |
| **PrEP outcomes** | | | | | | **RR (95% CI)** | ***p*** | **RR (95% CI)** | ***p*** | **RR (95% CI)** | ***p*** |
| Initiation *(primary)* | n | 142 | 670 | 700 | 79 | | | | | | |
| | Mean annual rate[2] | 3.9 | 12 | 19 | 1.8 | 1.78 (0.68–4.7) | 0.23 | 4.5 (1.23–16) | 0.024 | 6.9 (3.1–15) | <0.001* |
| Continuation *(primary)* | n | 27 | 270 | 243 | 31 | | | | | | |
| | Mean annual rate[2] | 0.73 | 4.8 | 6.6 | 0.72 | 0.88 (0.28–2.8) | 0.82 | 4.5 (0.93–22) | 0.061 | 5.8 (1.94–17) | 0.002* |
| **PEP outcomes** | | | | | | **RR (95% CI)** | ***p*** | **RR (95% CI)** | ***p*** | **RR (95% CI)** | ***p*** |
| Initiation | n | 491 | 950 | 925 | 172 | | | | | | |
| | Mean annual rate[2] | 13 | 17 | 25 | 4.0 | 3.6 (1.39–9.6) | 0.009* | 4.0 (1.39–12) | 0.011* | 5.8 (2.2–15) | <0.001* |
| Recurrent PEP use | n | 4 | 12 | 17 | 2 | | | | | | |
| | Mean annual rate[2] | 0.11 | 0.21 | 0.46 | 0.05 | 3.1 (0.35–27) | 0.31 | 4.8 (0.69–34) | 0.11 | 10 (1.43–70) | 0.021* |
| PEP-PrEP transition | n | 14 | 87 | 75 | 3 | | | | | | |
| | Mean annual rate[2] | 0.38 | 1.5 | 2.0 | 0.07 | 4.9 (1.16–21) | 0.031* | 16 (3.7–73) | <0.001* | 21 (5.7–76) | <0.001* |
| **PrEP or PEP outcomes** | | | | | | **RR (95% CI)** | ***p*** | **RR (95% CI)** | ***p*** | **RR (95% CI)** | ***p*** |
| Initiation | n | 618 | 1,532 | 1,549 | 248 | | | | | | |
| | Mean annual rate[2] | 17 | 27 | 42 | 5.8 | 2.9 (1.27–6.7) | 0.012* | 4.0 (1.43–11) | 0.009* | 6.0 (2.8–13) | <0.001* |
| Continuation or recurrent use | n | 46 | 357 | 331 | 36 | | | | | | |
| | Mean annual rate[2] | 1.3 | 6.4 | 8.9 | 0.84 | 1.33 (0.43–4.1) | 0.61 | 5.4 (1.25–23) | 0.024* | 7.1 (2.6–19) | <0.001* |

**Abbreviations:** Confidence interval (CI); Interquartile range (IQR); post-exposure prophylaxis (PEP); pre-exposure prophylaxis (PrEP); rate ratios (RR).

*Significant at Bonferroni–adjusted threshold of p=0.017 for primary outcomes of PrEP initiation and continuation. Significant at p=0.05 for all other outcomes.

[1]We estimated rate ratios using quasi-Poisson generalized linear models with a log link, modeling outcome counts with an offset for the total client volume and adjusting for county group. 95% CIs and p-values use HC3 robust standard errors. Missing outcomes were imputed as failures.

[2]Mean annual pharmacy rates were weighted by client volume and standardized to the median pharmacy volume in the clinic referral arm (2,030 clients/month).
